# Risk mitigating behaviours in people with inflammatory joint and skin disease during the COVID-19 pandemic differ by treatment type: a cross-sectional patient survey

**DOI:** 10.1101/2020.11.05.20226662

**Authors:** SK Mahil, M Yates, SM Langan, ZZN Yiu, T Tsakok, N Dand, KJ Mason, H McAteer, F Meynell, B Coker, A Vincent, D Urmston, A Vesty, J Kelly, C Lancelot, L Moorhead, H Bachelez, IN Bruce, F Capon, CR Contreras, AP Cope, C De La Cruz, P Di Meglio, P Gisondi, K Hyrich, D Jullien, J Lambert, H Waweru, H Marzo-Ortega, I McKinnes, L Naldi, S Norton, L Puig, R Sengupta, P Spuls, T Torres, RB Warren, J Weinman, CM Griffiths, JN Barker, MA Brown, JB Galloway, CH Smith, On behalf of the PsoProtect and CORE-UK study groups

**Author notes:** joint first authors (SK Mahil, M Yates).

## Abstract

**Objectives:** Registry data suggest that people with immune-mediated inflammatory diseases (IMIDs) receiving targeted systemic therapies have fewer adverse COVID-19 outcomes compared to patients receiving no systemic treatments. We used international patient survey data to explore the hypothesis that greater risk-mitigating behaviour in those receiving targeted therapies may account, at least in part, for this observation.

**Methods:** Online surveys were completed by individuals with Rheumatic and Musculoskeletal Diseases (RMD) (UK only) or psoriasis (globally) between 4th May and 7th September 2020. We used multiple logistic regression to assess the association between treatment type and risk-mitigating behaviour, adjusting for clinical and demographic characteristics. We characterised international variation in a mixed effects model.

**Results:** Of 3,720 participants (2,869 psoriasis, 851 RMD) from 74 countries, 2,262 (60.8%) reported the most stringent risk-mitigating behaviour (classified here under the umbrella term ‘shielding’). A greater proportion of those receiving targeted therapies (biologics and JAK inhibitors) reported shielding compared to those receiving no systemic therapy (adjusted odds ratio [OR] 1.63, 95% CI 1.35-1.97) and standard systemic agents (OR 1.39, 95% CI 1.22-1.56). Shielding was associated with established risk factors for severe COVID-19 (male sex [OR 1.14, 95% CI 1.05-1.24], obesity [OR 1.38, 95% CI 1.23-1.54], comorbidity burden [OR 1.43, 95% CI 1.15-1.78]), a primary indication of RMD (OR 1.37, 95% CI 1.27-1.48) and a positive anxiety or depression screen (OR 1.57, 95% CI 1.36-1.80). Modest differences in the proportion shielding were observed across nations.

**Conclusions:** Greater risk-mitigating behaviour among people with IMIDs receiving targeted therapies may contribute to the reported lower risk of adverse COVID-19 outcomes. The behaviour variation across treatment groups, IMIDs and nations reinforces the need for clear evidence-based patient communication on risk mitigation strategies and may help inform updated public health guidelines as the pandemic continues.

**Key messages:** *What is already known about this subject?:*

- At the beginning of the COVID-19 pandemic, patients with immune mediated inflammatory diseases (IMIDs) on targeted systemic immunosuppressive therapy were considered to be at higher risk of severe COVID-19. Subsequent registry data suggest that this may not the case.

*What does this study add?:*

- Here we characterise shielding behaviour in patients with IMIDs from a global survey. We identified that targeted systemic therapy associates with increased shielding behaviour, as do demographic risk factors for severe COVID-19 including male gender and obesity.
- Shielding behaviour varies across nations, albeit modestly when case-mix is taken into account.

*How might this impact on clinical practice or future developments?:*

- Variable shielding behaviour amongst patients with IMIDs may be an important confounder when considering differential COVID-19 risk between therapy types, so should be accounted for in analyses where possible.

## Introduction

The COVID-19 pandemic, caused by the highly infectious SARS-CoV-2 virus, represents an unprecedented global health crisis^1,2^. Death from COVID-19 is associated with male gender, older age, Asian/Black ethnicity, and coexisting conditions including cardiovascular disease and obesity^3,4^. Guided by international recommendations from the World Health Organization (WHO), public health risk mitigating measures such as social/physical distancing were introduced early in the pandemic to limit community transmission of COVID-19^5–8^. The WHO also recommended more stringent protection measures to reduce exposure risk in groups at higher risk of severe COVID-19^9^. This was referred to as ‘shielding’, and in the UK, was incorporated into Government policy where individuals classed as clinically extremely vulnerable were advised to physically isolate at home and avoid face-to-face interactions^7^.

Informed by pre-COVID-19 observational studies on drug-related risks of serious infection^10–13^, individuals with immune-mediated inflammatory diseases (IMIDs) receiving drugs that affect the immune system were considered at higher risk of severe COVID-19^14,15^. Whilst limited evidence has been published to date on drug-specific COVID-19 risks in IMIDs, rheumatoid arthritis, systemic lupus erythematosus and psoriasis were collectively suggested as risk factors for death using UK primary care data linked to hospital records from 17 million adults^3^. Global clinician-reported registry data in rheumatic diseases, psoriasis and inflammatory bowel disease have further suggested a differential risk of severe COVID-19 associated with different treatment types. In particular, use of targeted systemic therapies (biologics and Janus Kinase [JAK] inhibitors) was associated with a reduced risk of adverse COVID-19 outcomes, compared with no treatment or standard systemic agents^16–18^. It remains unclear if this is attributable to therapeutic modulation of the host antiviral immune and inflammatory response (i.e. biological mechanisms) or enhanced shielding behaviour in patients receiving targeted therapies (resulting in a lower infectious dose of SARS-CoV-2). There is an urgent need to address this knowledge gap since targeted and standard systemic therapies represent the mainstay of treatment in moderate to severe IMIDs.

Rheumatic and musculoskeletal diseases (RMDs) and psoriasis are common IMIDs that are closely related; psoriasis is the commonest immune-mediated skin disease associated with inflammatory arthritis, both conditions have a high prevalence of multimorbidity and are effectively treated with targeted and standard systemic therapies. We focused on RMDs and psoriasis as representative IMIDs and used global self-report survey data to explore the notion that individuals receiving different types of treatment exhibit distinct risk mitigating behaviours in the pandemic.

## Methods

### Study design, participants

Two online self-report surveys with aligned questions, permitting a combined analysis of data, were developed for people with psoriasis (Psoriasis Patient Registry for Outcomes, Therapy and Epidemiology of COVID-19 Infection *Me* [PsoProtect*Me*]; www.psoprotectme.org) and RMDs (COVID-19 Rheumatology Register [CORE-UK]; https://www.redcap02.medstats.org.uk/redcap/surveys/?s=LCA3L4JHXW). PsoProtect*Me* (available in 8 different languages) was promoted globally following its launch on 4th May 2020 and CORE-UK was subsequently launched on 12th June 2020 and promoted in the UK. The surveys were disseminated via social media, patient organisations (Table S1) and clinical networks. The eligibility criterion was any person (all ages) with a clinician-confirmed diagnosis of psoriasis (PsoProtect*Me*) or RMD (CORE-UK), irrespective of COVID-19 status (REC ref 20/YH/0135). Data were collected and managed using REDCap electronic data capture tools licensed to King’s College London Division of Health and Social Care Research^19^.

### Variables

Minimum sufficient core sets of variables within the surveys were defined by our study group of clinicians, epidemiologists, health data researchers and patient representatives. Patient Health Questionnaire-2 (PHQ-2) and Generalized Anxiety Disorder 2-item scale (GAD-2) were used to screen for depression and anxiety, respectively; scores of 3 or more were positive. Adherence was assessed with a single item question which asked if the individual had stopped or delayed their medication in the pandemic.

Risk mitigating behaviour was assessed with the following question: ‘*Over the past 30 days, what methods have you been using to protect yourself from COVID-19?’*. Respondents could select any of the following options: (1) Shielding (quarantine, strict distancing from family members in the home); (2) Self-isolation (quarantine, staying home, avoiding others); (3) Social distancing (avoiding crowds and large groups of people); (4) Using gloves and/or masks during social interactions; (5) None. The most stringent risk-mitigating behaviour was classified under the umbrella term ‘shielding’, encompassing (1) shielding and (2) self-isolation. Shielding was considered as a binary variable; any respondent who selected options (1) or (2) were coded as having shielded, and those selecting (3), (4) or (5) as having not shielded.

### Statistical methods

Data were extracted on 7th September 2020 and analyzed using Stata version 16. Continuous variables were reported using means and standard deviations (SD), and categorical/dichotomous variables as numbers and percentages. To account for partially completed surveys, respondents who completed more than 50% of variables were included. Individuals completing CORE-UK and PsoProtect*Me* were classified as having a primary diagnosis of RMD and psoriasis, respectively.

We characterised the demographic, socio-economic and disease-specific factors associated with the primary outcome of shielding behaviour in the pandemic. The key exposure measure was IMID treatment type in the pandemic, comprising 3 mutually exclusive categories:

#### (1) Targeted therapy: biologics and JAK inhibitors

(TNF inhibitors: adalimumab, certolizumab pegol, etanercept, infliximab, golimumab; IL-17 inhibitors: brodalumab, ixekizumab, secukinumab; IL12/IL-23p40 or IL-23p19 inhibitors: guselkumab, risankizumab, tildrakizumab, ustekinumab; IL-6 inhibitors tocilizumab, sarilumab; JAK inhibitors: baricitinib, tofacitinib);

#### (2) Standard systemic therapy

(methotrexate, ciclosporin, azathioprine, mycophenolate mofetil, fumaric acid esters/dimethylfumarate, sulfasalazine, leflunomide, acitretin, apremilast, chloroquine, hydroxychloroquine, prednisolone, tacrolimus);

#### (3) No systemic therapy

Patients on combination targeted and standard systemic therapy were included in the targeted therapy group, and surveys with missing treatment data were excluded. Apremilast was included in the standard systemic therapy group since in clinician-reported registry analyses, it was not grouped with biologics (unlike JAK inhibitors)^17,18^.

After excluding participants who self-reported COVID-19, associations with shielding status were assessed using: (a) a minimally adjusted logistic regression model including age and sex covariates; and (b) a fully adjusted model including a consensus list of covariates selected *a priori* as potentially influential on shielding behaviour on the basis of expert clinical opinion and existing evidence^20^. Treatment was included as a categorical variable in the fully adjusted model, with no systemic therapy as the reference group. Country of residence was included as a cluster variable.

Two sensitivity analyses were performed on the fully adjusted multivariable regression models: (1) Multiple imputation using iterative chained equations with 20 sets of imputed data to account for missing covariate data; (2) Exclusion of respondents on no systemic therapy, with standard systemic therapy becoming the reference group. Adherence data was included as a covariate in this model.

As the COVID-19 pandemic progressed in countries over different time periods, we hypothesised that the impact of time on the relationship between treatment and shielding behavior would vary between countries. To explore this, unadjusted estimates of shielding over time by treatment group were plotted for UK and non-UK survey respondents. Based on these plots we re-ran the multivariable model with UK respondents only including time as an interaction term with treatment, and as a fixed covariate, with a comparison of model fit. Time was converted to a binary variable, before or after 30th June. Shielding in the UK appeared to decrease after this date, which also coincided with the reopening of hospitality across the UK.

To characterise international variations in shielding behaviour, a mixed effects logistic regression model was executed with country of residence as a random effect. The random effect captures the difference between national and overall sample means, enabling estimation of case-mix adjusted rates. The national effects on shielding were visualised using a caterpillar plot^21^.

## Results

### Demographic, socio-economic and clinical characteristics of 3,720 study participants

Self-reported data from 3,720 individuals with a primary diagnosis of RMD (851, 22.9%) or psoriasis (2869, 77.1%) were available from 74 countries (including UK (2,578, 69.4%), Portugal (200, 5.4%), USA (165, 4.5%)) (demographic/clinical/socio-economic descriptions, Table 1). Two thousand two hundred and ninety-nine (61.8%) participants were not receiving a systemic agent for their psoriasis or RMD, 924 (24.8%) were receiving targeted therapies and 497 (13.4%) standard systemic agents. The three treatment groups had similar baseline characteristics including age, ethnicity, mean number per household and comorbidities. Non-adherence was also similar; 90 of 495 (18.2%) patients in the standard systemic therapy group reported non-adherence against medical advice, compared to 138 of 923 (15.0%) receiving targeted therapy. Of 257 (7.1%) participants who self-reported suspected or confirmed COVID-19, 80 (31.1%) were receiving a targeted therapy, 46 (17.9%) a standard systemic agent and 131 (51.0%) no systemic treatment. A lower proportion of those with COVID-19 reported shielding (143 of 257, 55.6%), compared to those without COVID-19 (2,051 of 3,352, 61.2%).

**Table 1.**
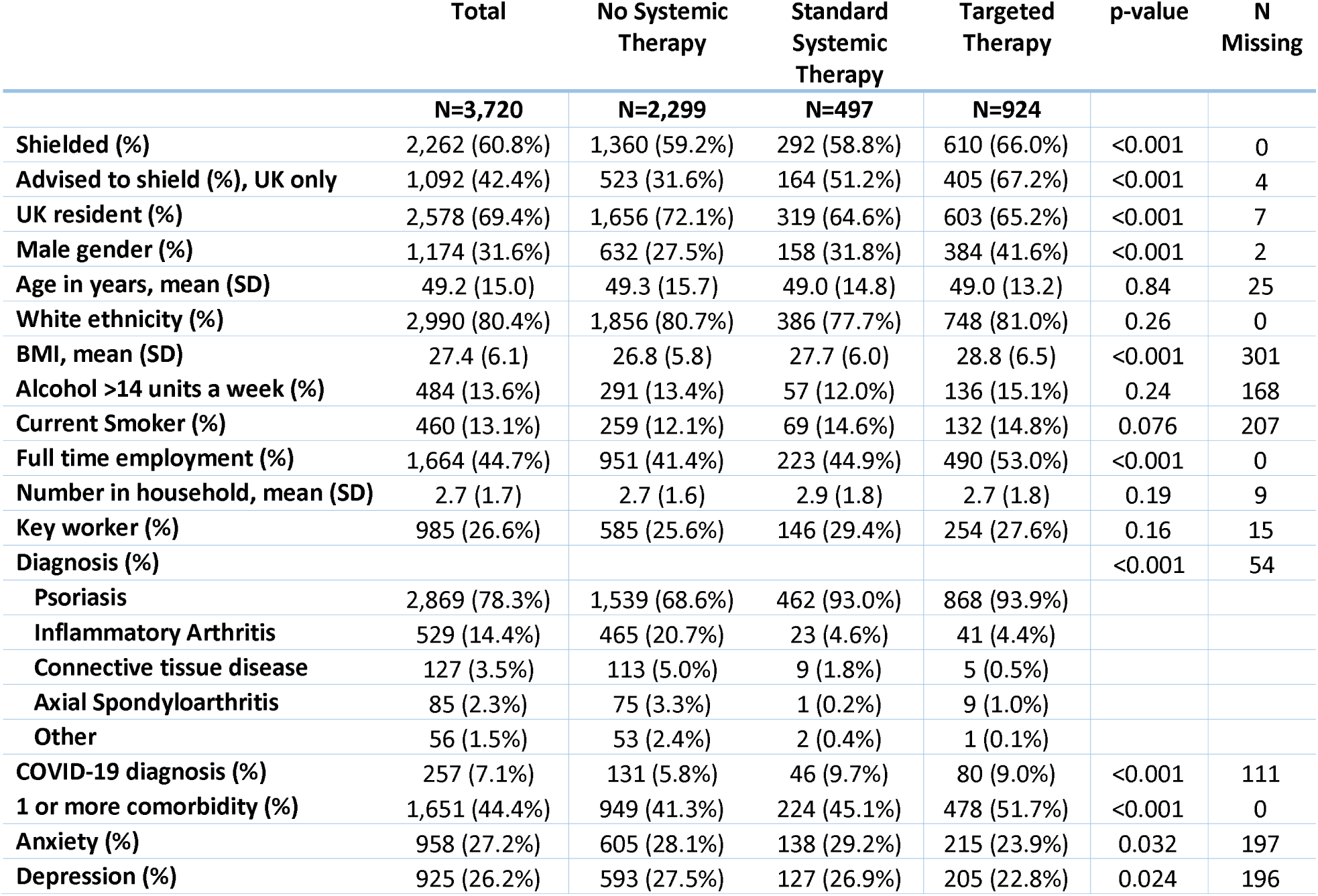
Participant characteristics, by treatment. ‘Shielded’ refers to participants who quarantined and self-isolated. Inflammatory arthritis included any participant with a diagnosis of rheumatoid arthritis or psoriatic arthritis. SD = standard deviation, BMI = body mass index, UK = United Kingdom.

### Risk mitigating behaviour differed by treatment type

Overall, 2,262 participants (60.8%) reported shielding. Of 1,632 participants self-reporting shielding in the UK, only 899 (55.1%) reported being specifically advised to shield (no data on shielding advice were available for non-UK participants). A greater proportion of those receiving targeted therapies (610 of 924, 66.0%) reported shielding compared with those receiving standard systemic agents (292 of 497, 58.8%) or no systemic therapy (1360 of 2299, 59.2%).

We used logistic regression models to investigate the observed differences in shielding by IMID treatment type. Compared to the reference group of no systemic therapy, an age and sex adjusted model for shielding behaviour estimated an odds ratio of 1.00 (95% CI 0.81 to 1.23) for those receiving standard systemic agents. In contrast, a significant association with shielding was observed for those receiving targeted therapies (OR 1.34, 95% CI 1.13 to 1.59) (Figure 1).

**Figure 1.**
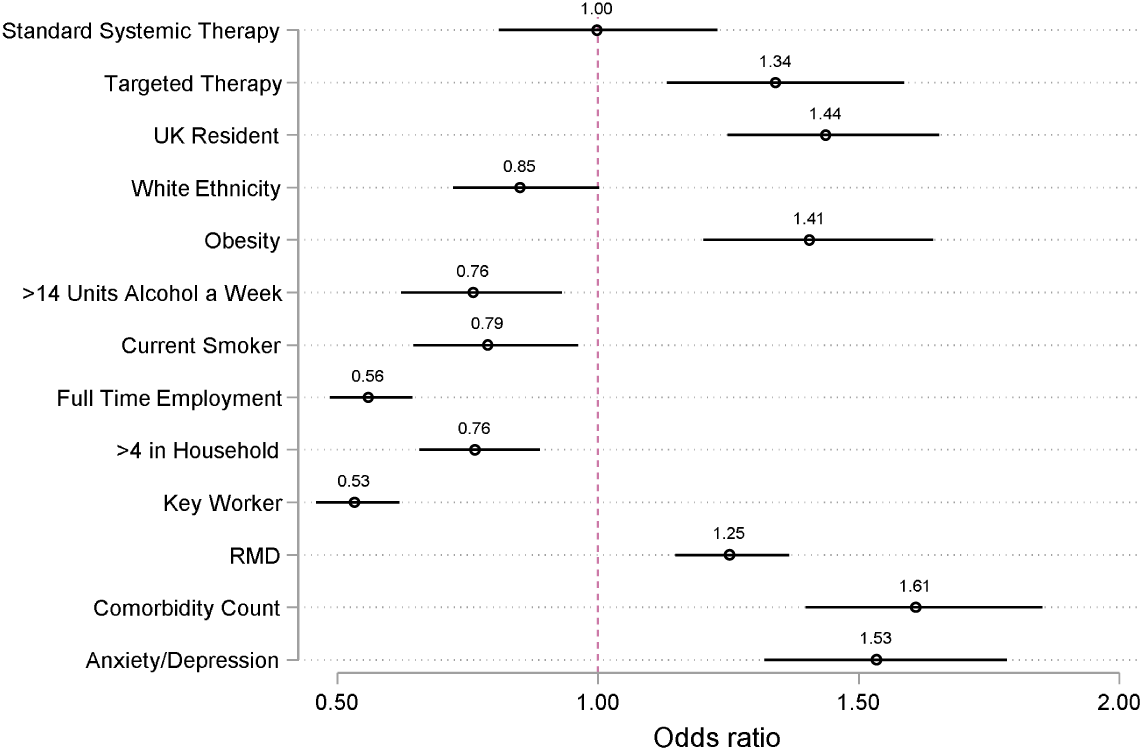
Age and gender adjusted associations with shielding. Each covariate was run as a predictor for shielding, adjusted for age and gender. Survey responders who were United Kingdom residents were asked if they received an NHS letter advising them to shield, which was associated with shielding, odds ratio of 4.7 (95% Confidence Interval 3.9 to 5.6). Standard therapy and biologic therapy were both compared to no systemic therapy as a reference group. UK = United Kingdom. RMD = rheumatic and musculoskeletal disease.

A fully adjusted multivariable logistic regression analysis was performed with a categorical treatment exposure variable: (1) targeted therapy; (2) standard systemic therapy; (3) no systemic therapy. The no systemic therapy group was used as the reference. Use of targeted therapy was associated with shielding compared to no systemic therapy (OR 1.63, 95% CI 1.35 to 1.97). Standard systemic therapy (OR 1.17, 95% CI 0.89 to 1.53) did not have a significant association with shielding. There were associations with shielding for RMD (OR 1.37, 95% CI 1.27 to 1.48), male sex (OR 1.14, 95% CI 1.05 to 1.24), comorbidity burden (OR 1.43, 95% CI 1.15 to 1.78), obesity (OR 1.37, 95% CI 1.23 to 1.54) and a positive anxiety or depression screen (OR 1.57, 95% CI 1.36 to 1.80) (Figure 2). In contrast, shielding was inversely associated with smoking (OR 0.73, 95% CI 0.63 to 0.85), full time employment (OR 0.66, 95% CI 0.49 to 0.88), >4 household members (OR 0.78, 95% CI 0.65 to 0.93) and key worker status (OR 0.55, 95% CI 0.44 to 0.68). No association was found with age (OR 1.00, 95% CI 0.99 to 1.00) or white ethnicity (OR 0.85, 95% CI 0.62 to 1.17).

**Figure 2.**
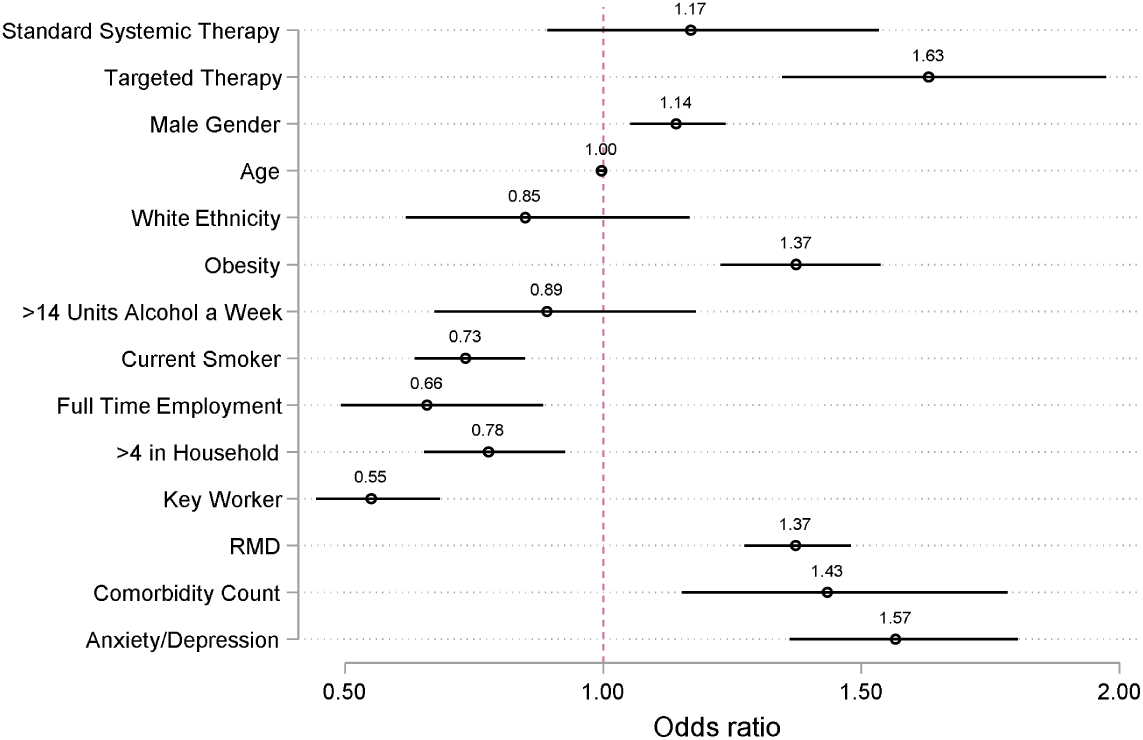
Fully adjusted model identifying associations with shielding. Covariates were determined a priori by an expert panel of collaborators. Country of residence was included as a cluster variable. Biologic therapy was compared to no systemic therapy as a reference group. RMD = rheumatic and musculoskeletal disease.

### Multivariable model sensitivity analyses

To account for missing data (Table 1), the multivariable model was rerun following multiple imputation. The magnitude and direction of associations did not change substantially (Table S2).

The model was also rerun excluding respondents on no systemic therapy, using standard systemic therapy as the reference group. The association between targeted therapy and shielding was preserved (OR 1.39, 95% CI 1.23 to 1.56). Therapy non-adherence was not associated with shielding (Table S3).

The influence of time (survey completion date) on shielding behaviour was explored across treatment groups. Estimated shielding behaviour generally decreased over time, however time had a differential impact in the UK (Figure S1) compared with non-UK countries (Figure S2). The multivariable model was therefore rerun with UK respondents only, first including time as a fixed covariate and secondly as an interaction term with treatment. The association between targeted therapy and shielding was preserved, with better model fit for the interaction term (further details in supplementary material).

### There was modest variation in risk mitigating behaviour across countries

A greater proportion of participants in the UK reported shielding compared to those elsewhere (63.3%, versus 55.0%). However, UK participants were also less likely to receive a targeted therapy (23.4%, versus 28.4%). A mixed effects model further showed modest variation around the sample mean in the proportion shielding in different countries, indicating broadly similar risk mitigating behaviours (Figure 3). Shielding was more prevalent in the UK, Canada and Argentina, but less prevalent in Portugal and Japan.

**Figure 3.**
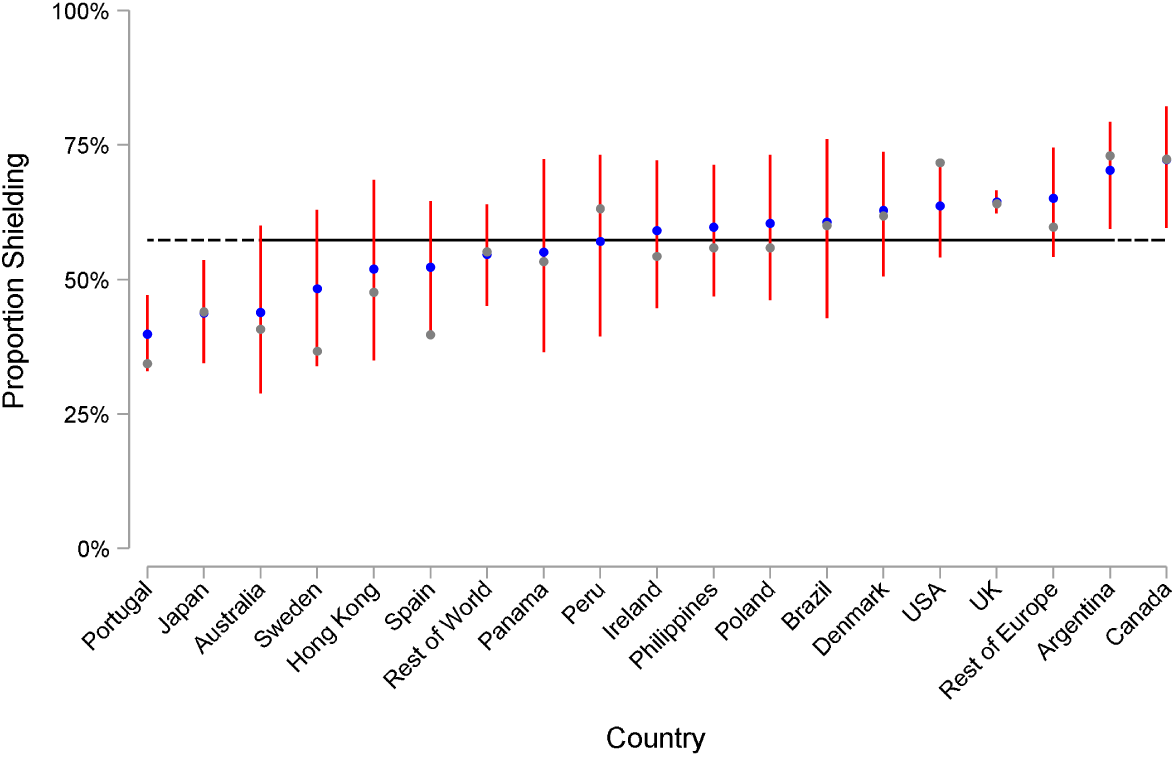
Caterpillar plot of observed and estimated risk mitigating behaviour, by nation. grey markers are the observed national proportions of survey respondents who shielded. The blue markers are the predicted random national effect on shielding from a mixed effects model, with 95% confidence intervals in red. The black horizontal line represents the overall mean. UK = United Kingdom, USA = United States of America.

## Discussion

We present global self-reported data on risk mitigating behaviours in 3,720 individuals with inflammatory joint and skin disease across 74 countries. Established risk factors for severe COVID-19 outcome including male sex, obesity and comorbidity burden were associated with stringent risk mitigating behaviour (classified here under the umbrella term ‘shielding’, encompassing self-reported shielding, quarantine, staying home or distancing within the home). Notably, use of targeted therapies (biologics and JAK inhibitors) was associated with shielding in comparison with no systemic therapy or standard systemic therapy. Although the differences in shielding behaviours across treatment groups in UK respondents were preserved when time was used as an interaction term, the observed decline in estimated shielding behaviour over time may help inform updated public health guidelines as the pandemic continues.

Our dataset is based on a large sample of individuals self-reporting RMD and psoriasis. Since there was no healthcare professional/record validation of survey responses, it is reassuring that key risk factors for severe COVID-19 in the general population such as male sex and obesity were associated with shielding. This is in keeping with public health messaging during the pandemic, and indicates a representative and generalizable sample. Shielding was recommended in groups of individuals deemed at higher risk^7,9^ on the premise that this would reduce the risk of COVID-19 transmission. More recently, evidence has emerged indicating shielding may also result in a less severe course of COVID-19 by reducing the frequency and intensity of exposures to SARS-CoV-2, thus lowering the infectious dose^22^. There is a growing body of evidence indicating that SARS-CoV-2 viral load positively correlates with disease severity^23,24^, and that in animal models, greater SARS-CoV-2 dose at exposure correlates with higher viral load and worse outcomes^25^.

Notably, increasing age was not associated with shielding behaviour in our dataset. This finding is in keeping with a recent international study of 8,317 individuals from the general population showing that age did not predict whether individuals took health precautions (mask wearing, social distancing, handwashing, staying home)^20^. Instead, beliefs that taking health precautions are effective and a concern for one’s own health were important predictors. Consistent with this, we identified an association between shielding and anxiety/depression. A larger proportion of participants also reported shielding compared to those advised to shield, which may reflect the elevated rates of self-reported anxiety. Anxiety and depression has also been reported in previous observational studies, underscoring the mental health burden of the pandemic (which may at least in part be due to the impact of social isolation)^26–28^. While this finding suggests accurate and representative data capture, more data are required on the severity and temporality of anxiety and depression.

Our study indicates a greater likelihood of shielding overall in individuals with a primary diagnosis of RMD compared with psoriasis, however the reasons underlying this are not clear. It may be attributable to differences in illness perception^29^, use of treatments and prevalence of comorbidities. IMID-specific COVID-19 risks are unknown, and neither RMD nor psoriasis were included in WHO and national public health shielding recommendations per se^7,9^. The reasons underlying differences in shielding behaviours between treatment groups, including patient perceptions of COVID-19 risk, also warrant further study. Although there is a paucity of data on treatment-related beliefs in psoriasis, recent single centre cross-sectional patient survey data in inflammatory bowel disease indicate patients perceive biologics to be riskier than other therapies^29^. These perceptions may influence shielding behaviours and are important to address during patient-clinician interactions.

Our global data on shielding behaviour builds on the findings from a recent single centre audit of 1,693 UK patients with rheumatic diseases^26^. Forty six percent self-reported shielding, however shielding among different treatment groups was not explored. In line with our findings, the audit found that a lower proportion of individuals with COVID-19 shielded (39%) compared to those without COVID-19 (47%). Our study also complements emerging findings from international clinician-reported registries, indicating differences in COVID-19 outcomes between different treatment types. Among 600 patients with rheumatic diseases and COVID-19 reported to the COVID-19 Global Rheumatology Alliance registry^17^, biologic/targeted synthetic systemic drug use was associated with lower odds of being hospitalized compared to patients receiving no systemic therapies. This effect was largely driven by TNF inhibitors since most patients on biologics were receiving this sub-group. A decreased risk of hospitalization or death was also associated with TNF inhibitor biologics compared with no treatment among 525 patients with inflammatory bowel disease and COVID-19 reported to SECURE-IBD^16^. In contrast, the standard systemics sulfasalazine or 5-aminosalicylate were associated with a higher risk of hospitalization or death. Our previously published study of 374 patients with psoriasis and COVID-19 reported to the PsoProtect registry further suggested an association between biologics (pooled data on TNF, IL-17 and IL-23 inhibitors) and reduced risk of hospitalization, compared to standard systemic therapies^18^. Although exploration of possible biological mechanisms underlying these associations is warranted (e.g. cytokine-targeted biologics may attenuate a severe systemic inflammatory response to COVID-19^30^), our current study highlights shielding behaviour as an important unmeasured potential mediator in these datasets. The differences in shielding behaviours across treatments supports the notion that greater protective shielding behaviour (resulting in a lower infectious dose of SARS-CoV-2) in those receiving targeted therapies may account, at least in part, for the observed associations. Thus conclusions from clinician-reported registry data about medication-related COVID-19 risk should be interpreted in this context, and further research efforts are required to quantify potential mediation through shielding.

A greater proportion of participants from the UK reported shielding compared with those elsewhere, which may reflect cross-national differences in public health messaging. However, these data should be interpreted with caution since our dataset is dominated by UK participants. Due to limited capture of socioeconomic data, we were unable to fully adjust for this confounder in the analysis, however we did identify that household density was inversely associated with shielding. Both shielding behaviour and clinical decision making around systemic therapies globally (including access to medications) may be influenced by socioeconomic variables such as income and education^31,32^, which may in turn influence outcome of COVID-19^4^. Linkage between health, social, behavioural and employment data should thus be prioritised in future research.

Collecting data via an online survey may have limited participation to more tech-literate individuals and those more connected to media. The study sample was mostly female (as expected in survey-based studies), of white ethnicity, and self-reported their diagnoses, which further limits the generalizability of the results. Ascertainment bias may overestimate the overall proportion shielding, since those more concerned about COVID-19 risk may be more likely to participate. Our sample, in which a greater proportion reported receiving targeted therapies compared with standard systemic agents, may not be representative of patients receiving systemic therapies more broadly. A disparate group of medications is also classified together as standard systemic agents. Potential selection bias may be addressed through systematic recruitment of participants enrolled in pharmacovigilance registries. Future linkage to registry and healthcare records may also validate self-reported demographic and clinical characteristics.

In the absence of a vaccine, shielding of at-risk individuals remains a global public health priority. Our study indicates that use of targeted therapies is associated with shielding in individuals with RMD and psoriasis, compared to no systemic treatment or standard systemic agents. This may contribute to the reported lower risk of adverse COVID-19 outcomes associated with targeted therapies reported by IMID registries. The observed differences in shielding across treatment groups, IMIDs, nations and time may inform future updates of public health recommendations for COVID-19 risk mitigating behaviours. Capture and consideration of risk mitigating behaviour is important in future studies of COVID-19 risk across people with IMIDs and on different types of systemic treatments.

## Data Availability

Summary level data available on reasonable request.

## Acknowledgements

We are very grateful to the patients who have contributed to PsoProtect*Me* and CORE-UK. We would like to acknowledge the professional and patient organizations who supported or promoted PsoProtect*Me* and CORE-UK (Table S1). We are grateful for the input of Prof Lars Iversen, Prof Nick Reynolds, Prof Joel Gelfand, Ms Christine Janus. We are also incredibly thankful to Engine Group UK for their generous creative input and website expertise.

## Funding

We acknowledge financial support from the Department of Health via the National Institute for Health Research (NIHR) Biomedical Research Centre based at Guy’s and St Thomas’ NHS Foundation Trust and King’s College London, the NIHR Manchester Biomedical Research Centre and the Psoriasis Association. The views expressed are those of the author(s) and not necessarily those of the NHS, the NIHR, or the Department of Health and Social Care. SKM is funded by a Medical Research Council (MRC) Clinical Academic Research Partnership award (MR/T02383X/1). ND is funded by Health Data Research UK (MR/S003126/1), which is funded by the UK MRC, Engineering and Physical Sciences Research Council; Economic and Social Research Council; Department of Health & Social Care (England); Chief Scientist Office of the Scottish Government Health and Social Care Directorates; Health and Social Care Research and Development Division (Welsh Government); Public Health Agency (Northern Ireland); British Heart Foundation; and Wellcome Trust. ZZNY is funded by a NIHR Academic Clinical Lectureship through the University of Manchester. CEMG is a NIHR Emeritus Senior Investigator and is funded in part by the MRC (MR/101 1808/1). CEMG and RBW are in part supported by the NIHR Manchester Biomedical Research Centre. SML is supported by a Wellcome senior research fellowship in clinical science (205039/Z/16/Z). SML is also supported by Health Data Research UK (grant no. LOND1), which is funded by the UK MRC, Engineering and Physical Sciences Research Council, Economic and Social Research Council, Department of Health and Social Care (England), Chief Scientist Office of the Scottish Government Health and Social Care Directorates, Health and Social Care Research and Development Division (Welsh Government), Public Health Agency (Northern Ireland), British Heart Foundation and Wellcome Trust.

## Competing Interests

Prof. Cope reports grants from Eli Lilly, outside the submitted work.

Dr. Vesty has nothing to disclose.

Dr. Vincent has nothing to disclose.

Mr. Bola Coker has nothing to disclose.

Dr. Capon reports consultancy fees from AnaptysBio, grants from Boheringer-Ingelheim, outside the submitted work;.

Dr De La Cruz has nothing to disclose.

Dr. Griffiths reports grants and personal fees from AbbVie, grants from Amgen, grants from BMS, grants and personal fees from Janssen, grants from LEO, grants and personal fees from Novartis, grants from Pfizer, grants from Almirall, grants and personal fees from Lilly, grants and personal fees from UCB Pharma, outside the submitted work;.

Dr. Contreras has nothing to disclose.

Dr. JULLIEN reports personal fees and non-financial support from Abbvie, personal fees and non-financial support from Novartis, personal fees and non-financial support from Janssen-Cilag, personal fees and non-financial support from Lilly, personal fees and non-financial support from Leo-Pharma, personal fees and non-financial support from MEDAC, personal fees and non-financial support from Celgene, personal fees from Amgen, outside the submitted work;.

Dr. Urmston reports grants from Almirral, grants from Abbvie, grants from Amgen, grants from Celgene, grants from Dermal Laboratories, grants from Eli Lilly, grants from Janssen, grants from LEO Pharma, grants from T and R Derma, grants from UCB, outside the submitted work;.

Dr. Meynall has nothing to disclose.

Dr. Bachelez reports personal fees from Abbvie, personal fees from Janssen, personal fees from LEO Pharma, personal fees from Novartis, personal fees from UCB, personal fees from Almirall, personal fees from Biocad, personal fees from Boehringer-Ingelheim, personal fees from Kyowa Kirin, personal fees from Pfizer, outside the submitted work;.

Dr. McAteer reports grants from Abbvie, grants from Almirral, grants from Amgen, grants from Celgene, grants from Dermal Laboratories, grants from Eli Lilly, grants from Janssen, grants from LEO Pharma, grants from UCB, grants from T and R Derma, outside the submitted work;.

Dr. Bruce reports grants from National Institute for Health Research, during the conduct of the study; grants from Genzyme Sanofi, grants and personal fees from GSK, grants and personal fees from UCB, personal fees from Eli Lilly, personal fees from Astra Zeneca, personal fees from MErck Serono, personal fees from Aurinia, personal fees from ILTOO, outside the submitted work;.

Dr. Barker reports grants and personal fees from Abbvie, grants and personal fees from Novartis, grants and personal fees from Lilly, grants and personal fees from J&J, from null, during the conduct of the study;.

Dr. Galloway reports personal fees from Abbvie, personal fees from Sanofi, personal fees from Novartis, personal fees from Pfizer, grants from Eli Lilly, personal fees from Janssen, personal fees from UCB, outside the submitted work;.

Dr. Lambert has nothing to disclose.

Prof Weinmann has presented talks for Abbvie, Abbott, Bayer, Chiesi, Boehringer Ingelheim, Roche and Merck.

Prof. Hyrich reports grants from Bristol Myers Squibb, grants from Pfizer, personal fees from Abbvie, outside the submitted work;.

Dr. MASON reports personal fees from LEO Pharma and Novartis, outside the submitted work.

Dr. Moorhead reports personal fees from Abbvie, personal fees from Celgene, personal fees from Janssen, personal fees from LEO Pharma, personal fees from Novartis, personal fees from UCB, outside the submitted work;.

Dr. Naldi has nothing to disclose.

Dr. Puig reports grants and personal fees from AbbVie, grants and personal fees from Almirall, grants and personal fees from Amgen, grants and personal fees from Boehringer Ingelheim, personal fees from Bristol Myers Squibb, personal fees from Fresenius-Kabi, grants and personal fees from Janssen, grants and personal fees from Lilly, personal fees from Mylan, grants and personal fees from Novartis, personal fees from Pfizer, personal fees from Sandoz, personal fees from Sanofi, personal fees from Samsung-Bioepis, grants and personal fees from UCB, outside the submitted work;.

Dr. Mahil reports grants from Abbvie, grants from Celgene, grants from Eli Lilly, grants from Janssen-Cilag, grants from Novartis, grants from Sanofi, from UCB|, outside the submitted work;.

Dr. Marzo-Ortega reports grants and personal fees from Janssen, grants and personal fees from Novartis, personal fees from Abbvie, personal fees from Celgene, personal fees from Eli Lilly, personal fees from Pfizer, personal fees from Takeda, personal fees from UCB, outside the submitted work;.

Prof. BROWN has nothing to disclose.

Prof. McInnes reports grants from Abbvie, grants from Astra Zeneca, grants from Bristol Myers Squibb, grants from Boerhinger, grants from Celgene, grants from Roche, grants from Janssen, grants from Novartis, grants from Eli Lilly, grants from UCB, outside the submitted work;.

Dr. Yates has nothing to disclose.

Dr. Dand has nothing to disclose.

Dr. Gisondi reports personal fees from Abbvie, Amgen, Eli Lilly, Jannsen, Novartis, Pierre Fabre, Sandoz, UCB, outside the submitted work;.

Dr. Di Meglio reports grants and personal fees from UCB, personal fees from Novartis, personal fees from Janssen, outside the submitted work;.

Dr Sengupta: Speaker fees, consultancy and/or grants from Abbvie, Biogen, Celgene, MSD, Novartis and UCB.

Dr. Warren reports grants and personal fees from Abbvie, grants and personal fees from Celgene, grants and personal fees from Eli Lilly, grants and personal fees from Novartis, personal fees from Sanofi, grants and personal fees from UCB|, grants and personal fees from Almirall, grants and personal fees from Amgen, grants and personal fees from Janssen, grants and personal fees from Leo, grants and personal fees from Pfizer, personal fees from Arena, personal fees from Avillion, personal fees from Bristol Myers Squibb, personal fees from Boehringer Ingelheim, outside the submitted work;.

Dr. Langan has nothing to disclose.

Dr. Smith reports grants from Abbvie, grants from Sanofi, grants from Novartis, grants from Pfizer, outside the submitted work;.

Dr. Norton has nothing to disclose.

Dr Spuls is a member of the ISAC of psoprotect.

Dr. Tsakok has nothing to disclose.

Dr. Torres reports grants and personal fees from AbbVie, Almirall, Amgen, Arena Pharmaceuticals, Biogen, Biocad, Boehringer Ingelheim, Bristol-Myers Squibb, Celgene, Eli Lilly, Janssen, LEO Pharma, MSD, Novartis, Pfizer, Samsung-Bioepis, Sandoz, during the conduct of the study.

Dr. Yiu has nothing to disclose.

Dr Waweru is on the Board of the International Federation of Psoriasis Associations who have received grants from Abbvie, Almirall, Amgen, Bristol Meyers Squibb, Boehringer Ingelheim, Celgene, Janssen, Leo Pharma, Eli Lilly, Novartis, Sun Pharma, Pfizer, and UCB, outside the submitted work;.

## Ethical Approval

Research approved by KCL research ethics committee (REC ref 20/YH/0135)

## Data Sharing

Summary level data available on reasonable request.

## Notes

### Author Declarations

Research approved by KCL research ethics committee (REC ref 20/YH/0135)

