## Supplementary material for "Risk mitigating behaviours in people with inflammatory joint and skin disease during the COVID-19 pandemic differ by treatment type: a cross-sectional patient survey"

| **Supplementary table 1. - Organisations who supported or promoted PsoProtect*Me* and/or CORE-UK** |
| --- |
| **Organization** |
| Psoriasis Association |
| European Society for Dermatological Research (ESDR) |
| International Psoriasis Council (IPC) |
| American Academy of Dermatology (AAD) |
| International Federation of Psoriasis Associations (IFPA) |
| Global Psoriasis Atlas (GPA) |
| Global Skin |
| National Psoriasis Foundation (NPF) |
| European Dermatology Forum (EDF) |
| International League of Dermatological Societies (ILDS) |
| British Association of Dermatologists (BAD) |
| Skin Inflammation and Psoriasis International Network (SPIN) |
| British Society for Investigative Dermatology (BSID) |
| European Academy of Dermatology and Venereology (EADV) |
| Irish Skin Foundation (ISF) |
| Psoriasis and Psoriatic Arthritis Alliance (PAPAA) |
| European Rare and Severe Psoriasis Expert Network (ERASPEN) |
| PSONET |
| British Skin Foundation (BSF) |
| European Umbrella Organisation for Psoriasis Movements (EUROPSO) |
| British Dermatological Nursing Group (BDNG) |
| Australasian Psoriasis Registry (APR) |
| Canadian Psoriasis Network (CPN) |
| French Psoriasis Research Group (Group de Recherche sur le Psoriasis, GRPSO) |
| Canadian Association of Psoriasis Patients (CAPP) |
| Amicus Foundation Psoriasis and PsA (Poland) |
| Civil Association for Psoriasis Patients (Asociación Civil para el Enfermo de Psoriasis, AEPSO, Argentina) |
| Danish Psoriasis Association (Psoriasisforeningen) |
| Finnish Psoriasis Association (Psoriasisliitto) |
| France Psoriasis |
| Fundación de Apoyo a Pacientes con Psoriasis (FUNAPAPSO, Dominican Republic) |
| Global Healthy Living Foundation (GHLF) |
| Hong Kong Psoriasis Patients Association |
| Japan Psoriasis Association (Inspire Japan WPD) |
| Psoriasis Action (Acción Psoriasis) |
| Psoriasis Association of Singapore |
| Psoriasis Group of the Spanish Academy of Dermatology and Venereology |
| Psoriasis New Life Association from El Salvador (Asociacion Psoriasis Nueva Vida El Salvador, PSONUVES) |
| Psoriasis of Panama Foundation (Fundacion Psoriasis de Panama) |
| Psoriasis Philippines (PsorPhil) |
| PsorViet (Vietnam) |
| Psychodermatology UK |
| Puerto Rican Association for Helping Psoriasis Patients (Asociacion Puertorriquena de Ayuda al Paciente de Psoriasis, APAPP) |
| Swedish Psoriasis Association (Psoriasisforbundet) |
| Union of Psoriasis and PsA Associations (Poland) |
| Uruguay Psoriasis Association (Asociación Psoriasis Uruguay, APSUR) |
| Venezuelan Association of Psoriasis (Asociación Venezolana de Psicología Social, AVEPSO) |
| British Association of Dermatologists Biologics Interventions Register (BADBIR) |
| Global Rheumatology Alliance |
| NCD Alliance |
| SECURE-AD |
| SECURE-Alopecia |
| SECURE-IBD  National Rheumatoid Arthritis Society  British Society for Rheumatology  British Society for SpondyloArthritis |

**Supplementary table 2. Imputed multivariable logistic regression model characterising association between therapy and shielding behaviour (reference group is no systemic therapy).**

| **Shielding** | **Odds Ratio** | **P value** | **95% Confidence Interval** |
| --- | --- | --- | --- |
| Targeted therapy | 1.64 | <0.001 | 1.35, 1.98 |
| Standard systemic therapy | 1.23 | 0.16 | 0.92, 1.64 |
| Age | 1.00 | 0.25 | 0.99, 1.01 |
| Male gender | 1.09 | 0.12 | 0.98, 1.22 |
| Comorbidity | 1.47 | <0.001 | 1.22, 1.79 |
| Alcohol intake | 0.86 | 0.31 | 0.65, 1.15 |
| Anxiety/Depression | 1.53 | <0.001 | 1.32, 1.78 |
| BMI | 1.03 | <0.001 | 1.01, 1.04 |
| Current smoker | 0.79 | <0.001 | 0.69, 0.99 |
| White ethnicity | 0.79 | 0.15 | 0.57, 1.09 |
| Full time employment | 0.62 | <0.001 | 0.48, 0.80 |
| Key worker | 0.58 | <0.001 | 0.48, 0.71 |
| RMD diagnosis | 1.34 | <0.001 | 1.24, 1.45 |
| Household density | 0.77 | 0.01 | 0.64, 0.93 |

**Supplementary table 3. Multivariable logistic regression model characterising association between biologic therapy and shielding behaviour with standard systemic therapy as the comparator group.**

| **Shielding** | **Odds Ratio** | **P value** | **95% Confidence Interval** |
| --- | --- | --- | --- |
| Targeted therapy | 1.39 | <0.001 | 1.23, 1.56 |
| Non-adherent to therapy | 1.01 | 0.9 | 0.66, 1.55 |
| Age | 0.99 | 0.09 | 0.99, 1.00 |
| Male gender | 1.12 | 0.16 | 0.96, 1.31 |
| Comorbidity | 1.47 | 0.04 | 1.02, 2.13 |
| Alcohol intake | 1.14 | 0.54 | 0.76, 1.70 |
| Anxiety/Depression | 1.49 | <0.001 | 1.18, 1.88 |
| BMI | 1.51 | <0.001 | 1.31, 1.75 |
| Current smoker | 0.60 | <0.001 | 0.43, 0.83 |
| White ethnicity | 1.00 | 0.99 | 0.69, 1.47 |
| Full time employment | 0.59 | 0.002 | 0.42, 0.83 |
| Key worker | 0.67 | <0.001 | 0.54, 0.83 |
| RMD diagnosis | 1.31 | 0.11 | 0.94, 1.82 |
| Household density | 0.66 | <0.001 | 0.57, 0.76 |

**Supplementary table 4. UK only analysis: multivariable logistic regression model characterising association between therapy and shielding behaviour (reference group is no systemic therapy).**

| **Shielding** | **Odds Ratio** | **P value** | **95% Confidence Interval** |
| --- | --- | --- | --- |
| Targeted therapy | 1.71 | <0.001 | 1.35, 2.15 |
| Standard systemic therapy | 1.33 | 0.05 | 1.00, 1.76 |
| Age | 1.00 | 0.7 | 0.99, 1.01 |
| Male gender | 1.14 | 0.22 | 0.93, 1.39 |
| Comorbidity | 1.65 | <0.001 | 1.36, 2.00 |
| Alcohol intake | 0.77 | 0.05 | 0.60, 0.99 |
| Anxiety/Depression | 1.68 | <0.001 | 1.37, 2.06 |
| BMI | 1.43 | <0.001 | 1.16, 1.76 |
| Current smoker | 0.74 | 0.04 | 0.56, 0.99 |
| White ethnicity | 0.64 | 0.01 | 0.46, 0.88 |
| Full time employment | 0.80 | 0.03 | 0.65, 0.98 |
| Key worker | 0.50 | <0.001 | 0.40, 0.62 |
| RMD diagnosis | 1.26 | <0.001 | 1.13, 1.40 |
| Household density | 0.86 | 0.18 | 0.70, 1.07 |

**Supplementary table 5. UK only analysis: multivariable logistic regression model characterising association between therapy and shielding behaviour (reference group is no systemic therapy), with time of survey completion included as a fixed covariate.**

| **Shielding** | **Odds Ratio** | **P value** | **95% Confidence Interval** |
| --- | --- | --- | --- |
| Targeted therapy | 1.70 | <0.001 | 1.34, 2.16 |
| Standard systemic therapy | 1.24 | 0.15 | 0.93, 1.65 |
| Survey completed After June 31^st^ 2020 | 0.41 | <0.001 | 0.33, 0.51 |
| Age | 1.00 | 0.60 | 0.99, 1.01 |
| Male gender | 1.15 | 0.20 | 0.93, 1.41 |
| Comorbidity | 1.71 | <0.001 | 1.41, 2.08 |
| Alcohol intake | 0.78 | 0.06 | 0.61, 1.01 |
| Anxiety/Depression | 1.74 | <0.001 | 1.41, 2.15 |
| BMI | 1.46 | <0.001 | 1.18, 1.81 |
| Current smoker | 0.76 | 0.06 | 0.57, 1.01 |
| White ethnicity | 0.59 | <0.001 | 0.42, 0.82 |
| Full time employment | 0.81 | 0.05 | 0.65 0.99 |
| Key worker | 0.49 | <0.001 | 0.39, 0.61 |
| RMD diagnosis | 1.62 | <0.001 | 1.42, 1.83 |
| Household density | 0.41 | <0.001 | 0.33, 0.51 |

**Supplementary table 6. UK only analysis: multivariable logistic regression model characterising association between therapy and shielding behaviour (reference group is no systemic therapy), with time of survey completion included as an interaction term with treatment.**

| **Shielding** | **Odds Ratio** | **P value** | **95% Confidence Interval** |
| --- | --- | --- | --- |
| Targeted therapy | 1.82 | <0.001 | 1.34, 2.48 |
| Standard systemic therapy | 1.35 | 0.1 | 0.95, 1.93 |
| 1.Survey completed After June 31^st^ 2020 | 0.44 | <0.001 | 0.34, 0.58 |
| Treatment#Survey completion time |  |  |  |
| Standard Systemic Therapy#1 | 0.78 | 0.43 | 0.43, 1.43 |
| Biologic Therapy#1 | 0.86 | 0.51 | 0.54, 1.36 |
| Age | 1.00 | 0.59 | 0.99, 1.01 |
| Male gender | 1.14 | 0.21 | 0.93, 1.41 |
| Comorbidity | 1.71 | <0.001 | 1.41, 2.08 |
| Alcohol intake | 0.78 | 0.06 | 0.61, 1.01 |
| Anxiety/Depression | 1.74 | <0.001 | 1.41, 2.15 |
| BMI | 1.46 | <0.001 | 1.18, 1.81 |
| Current smoker | 0.76 | 0.07 | 0.57, 1.02 |
| White ethnicity | 0.59 | <0.001 | 0.42, 0.83 |
| Full time employment | 0.81 | 0.05 | 0.65, 1.00 |
| Key worker | 0.49 | <0.001 | 0.39, 0.61 |
| RMD diagnosis | 1.59 | <0.001 | 1.40, 1.81 |
| Household density | 0.89 | 0.29 | 0.72, 1.11 |

**Supplementary figure 1. Estimated shielding over time, UK respondents only.**

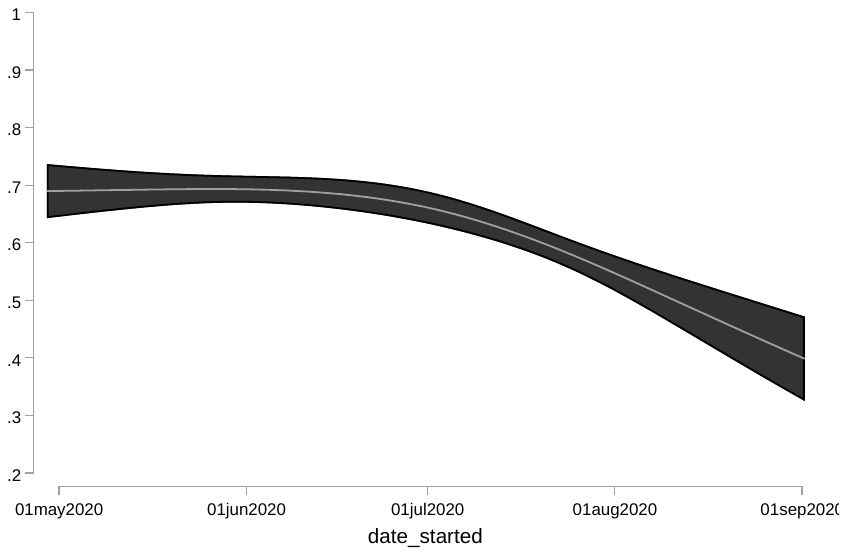

**Supplementary figure 1**. Shielding behaviour over time was estimated via logistic regression, with time converted to a cubic spline with three knots. The black shaded areas indicate 95% confidence intervals.

**Supplementary figure 2. Estimated shielding over time, non-UK respondents only.**

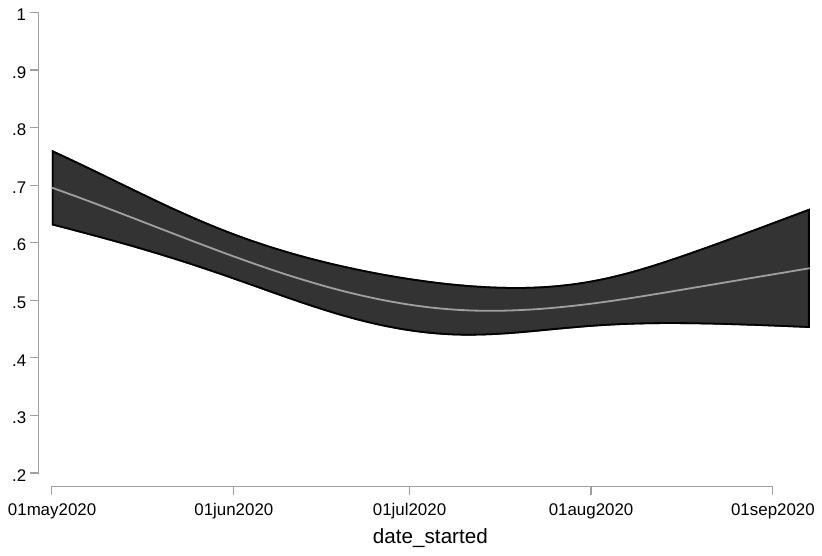

**Supplementary figure 2.** Shielding behaviour over time was estimated via logistic regression, with time converted to a cubic spline with three knots. The black shaded areas indicate 95% confidence intervals.

**Supplementary table 7. Therapy breakdown.**

| **Therapy** | **Frequency** |
| --- | --- |
| Acitretin | 49 |
| Apremilast | 64 |
| Ciclosporin | 41 |
| Dexamethasone | 1 |
| Fumaric Acid | 12 |
| Methotrexate_tab | 317 |
| Prednisolone | 14 |
| Brodalimumab | 9 |
| Guselkumab | 48 |
| Ixekizumab | 66 |
| Methotrexate_inj | 78 |
| Risankizumab | 14 |
| Secukinumab | 136 |
| Tildrakizumab | 2 |
| IL-12/23 Inhibitors | 194 |
| Abatacept | 1 |
| Hydroxychloroquine | 16 |
| IL-6 Inhibitors | 3 |
| JAK Inhibitors | 4 |
| Mycophenolate | 1 |
| Rituximab | 1 |
| Sulphasalazine | 5 |
| TNF Inhibitors | 345 |
